# Effects of cardiovascular disease risk factors on cardiorespiratory fitness in firefighters – a protocol for a systematic review and meta-analysis

**DOI:** 10.1101/2022.08.29.22279323

**Authors:** Jaron Ras, Elpidoforos S. Soteriades, Andre P. Kengne, Denise Smith, Lloyd Leach

**Author notes:** Corresponding author: Department of Sport, Recreation and Exercise Science, Faculty of Community and Health Sciences, University of the Western Cape, Cape Town, South Africa. Jaron Ras.

## Abstract

**Introduction:** Globally, firefighting is recognized as among the most physically demanding professions. In addition, because of the hazardous nature of firefighting, firefighters are required to wear heavy insulated personnel protective equipment that protects them from exposures to hazardous chemicals and fumes and severe temperatures. This requires that, throughout their careers, firefighters maintain their cardiovascular health and maintain a satisfactory cardiovascular fitness to manage these stressors and perform their duties sufficient vigour. The aim of this systematic is to determine the effects of cardiovascular disease risk factors on cardiorespiratory fitness of firefighters.

**Methods:** Using the comprehensive search terms, a detailed literature search, with no limitation to publication year, will be conducted to identify relevant studies from PubMed/Medline, Web of Science, EBSCOHost, ScienceDirect and SCOPUS. Data-extraction will be extracted using a researcher-generated data extraction form. Data will be imported into Zotero® version 6.0.9, and duplicates removed. The article screening will be conducted using the Rayyan® intelligent systematic review tool. Thereafter, information from the included studies will be captured on the researcher-generated data extraction form. For the methodological assessment, the Appraisal Tool for Cross Sectional Studies (AXIS) checklist and the Critical Appraisal Skills Programme (CASP) toolkit will be used. For the meta-analysis, Review Manager 5.3 will be used to determine the exposure effects and MedCalc® statistical software Ltd. to determine the pooled correlation effects.

**Discussion:** This systematic review arose from the global cardiovascular health concerns that firefighters face, and how the development and progression of CVD risk factors effect cardiorespiratory fitness in firefighters. Cardiorespiratory has been shown to be an essential factor in optimal occupational performance in firefighters but understudied related to CVD risk factors. It is anticipated that this review will make a considerable contribution to the international scientific literature on the effect that CVD risk factors will have on cardiorespiratory fitness.

**Systematic review protocol registration:** PROSPERO (CRD42021258898)

## INTRODUCTION

Globally, firefighting is recognized as among the most physically demanding professions [1– 4]. In addition, because of the hazardous nature of firefighting, firefighters are required to wear heavy insulated personnel protective equipment that protects them from exposures to hazardous chemicals and fumes and severe temperatures [1–4]. The full PPE worn with the self-contained breathing apparatus (SCBA) may weigh up to 29.3 kg, which places significant strain on the cardiovascular system [1]. It has been shown that firefighters routinely exceed their age predicted maximal heart rates while performing work-related duties [5–7], which significantly predisposes firefighters to sudden cardiac events while on duty [3,4,8], which may account for the high mortality rates in this population.

Studies have shown that numerous firefighters have multiple cardiovascular disease (CVD) risk factors, with the most prevalent risk factors being obesity, cigarette smoking, dyslipidemia, and physical inactivity [9–15]. This is a concern given the physically strenuous occupation of firefighting [2–4,8]. Numerous studies have reported that firefighters have an inadmissibly high percentage of mortality due to cardiovascular related incidents, with the preponderance of these events related to underlying CVD risk factors [4,8,16]. An additional factor contributing toward the high prevalence of cardiovascular-related mortality is inadequate physical fitness, which results in increased cardiovascular strain and overexertion [17,18]. Firefighters are, therefore, required to remain in peak cardiorespiratory conditioning, while maintaining good CVD health throughout their careers [4,8,17]. Firefighters that have multiple CVD risk factors, and have suboptimal cardiorespiratory conditioning, are not only at risk for sustaining a SCE while on duty, but also at risk for endangering the public they serve to protect [4,8,16,17].

This study arose from the difficulties that firefighters routinely encounter, both globally and, in particular, locally in South Africa [12,19]. A disconcerting quantity of firefighters are at increased CVD risk and have poor cardiorespiratory fitness levels, which negatively impacts their overall health, well-being, and occupational performance [20–24]. The development of these CVD risk factors diminish firefighters ability to cope with the physical strain of firefighting, that has been described as being comparable to the demands placed on elite sportspersons [25]. It has been shown, in previous systematic reviews, that CVD risk factors are significantly related to cardiorespiratory fitness [26–29]. However, to the best of the authors knowledge, no systematic review and meta-analysis has investigated the effect of CVD risk factors on cardiorespiratory fitness in firefighters, which motivated the current systematic review. One of the intentions of this review was to assist in informing policy change in South Africa, specifically on the need for remedial action and the development of strategies to enhance and maintain the cardiorespiratory fitness levels and cardiovascular wellbeing of firefighters.

## METHODS AND ANALYSIS

### Study Design

This protocol was registered in the PROSPERO (International prospective register of systematic reviews), with the registration number CRD42022330510. The systematic review protocol is reported in accordance with the Preferred Reporting Items for Systematic Review and Meta-Analysis Protocol (PRISMA-P). The systematic review methodology will be conducted in accordance with the guidelines Meta-analysis of Observational Studies in Epidemiology studies (MOOSE) and Quality of Reporting of Meta-analysis (QUOROM) [30,31]. When considering studies for this review, the PRISMA guidelines will be followed.

The authors have chosen to investigate the effect of CVD risk factors on cardiorespiratory fitness in adult, full-time firefighters. The focus of this review will be to determine the effect of CVD risk factors on cardiorespiratory fitness of firefighters. A quantitative systematic review is the preferred study design for this review [32].

### Aim of the Review

The aim of this systematic review is to determine the effects of cardiovascular disease risk factors on the cardiorespiratory fitness of firefighters.

### Research Question

What effects do cardiovascular disease risk factors have on the cardiorespiratory fitness of firefighters?

### Objectives of the Review

The objective of the study is to investigate the effects of cardiovascular disease risk factors on the cardiorespiratory fitness of firefighters.

### Participants

Full-time, part-time and volunteer firefighters 18 years and older.

### Exposure Types

Cardiovascular disease risk factors in relation to the cardiorespiratory fitness of firefighters.

### Outcome Types

i. Cardiovascular disease risk factors related to the cardiorespiratory fitness of firefighters.

### Search Strategy for Identification of Studies

Using the comprehensive search terms, a detailed literature search will be conducted to identify studies investigating the effects of cardiovascular disease risk factors on the cardiorespiratory fitness of firefighters. Details on the inclusion and exclusion criteria for the systematic review is contained in Table 1:

**Table 1:** Inclusion and exclusion criteria of studies.

| Inclusion Criteria | Exclusion Criteria |
| --- | --- |
| i. Studies that included full-time, part-time and volunteer adult firefighters between the ages of 18 and 65 years, with no limitations to publication year. | i. Studies not focusing on cardiorespiratory fitness as the outcomes. |
| ii. Observational studies, such as cross-sectional, cohort and case controlled, and experimental study designs. | ii. Studies not focusing on cardiovascular disease risk factors as the main exposures. |
| iii. Studies investigating the effects of cardiovascular disease risk factors on cardiorespiratory fitness in firefighters. | iii. Systematic reviews or other types of reviews. |
|  | iv. Articles that are non-English. |

The team of reviewers responsible for study identification will be made up of three main contributors:

i. Reviewer I (JR) will be the primary investigator. JR will assume responsibility for all aspects of the review and will be one of the reviewers tasked with conducting the literature search, independently extracting the data, responsible for verifying the data collected, analysing the results, grading the quality of the data, and write up of the first draft of the systematic review.
ii. Reviewer II (RN) will be the second reviewer and be tasked with independently conducting a literature search, extracting the data of studies, validating the data collected, and grading the quality of the data.
iii. Reviewer III (LL) will serve as the adjudicator and resolve any disagreements between the two independent reviewers.

#### Electronic Literature Search

Using the comprehensive search terms, an extensive literature search will be conducted to enable the capturing of as many relevant articles as possible to be included in the systematic review and meta-analysis, however, these studies will be limited to English papers only. The academic journal databases that will be searched will include PubMed/Medline, Web of Science, SCOPUS, ScienceDirect and EBSCOhost. The literature search will take place between August and September 2022. In addition, there will be no limitation to publication year to allow inclusivity. The comprehensive search strategy will include the arrangement of keywords and medical subject heading (MeSH) in various arrangements depending on the specific database searched. In Table 2, the various arrangements of each of the search terms for the population and the cardiovascular disease risk factors will be combined with each of the outcome terms. The search strategies for the other databases are presented in Appendix 1.

**Table 2:**
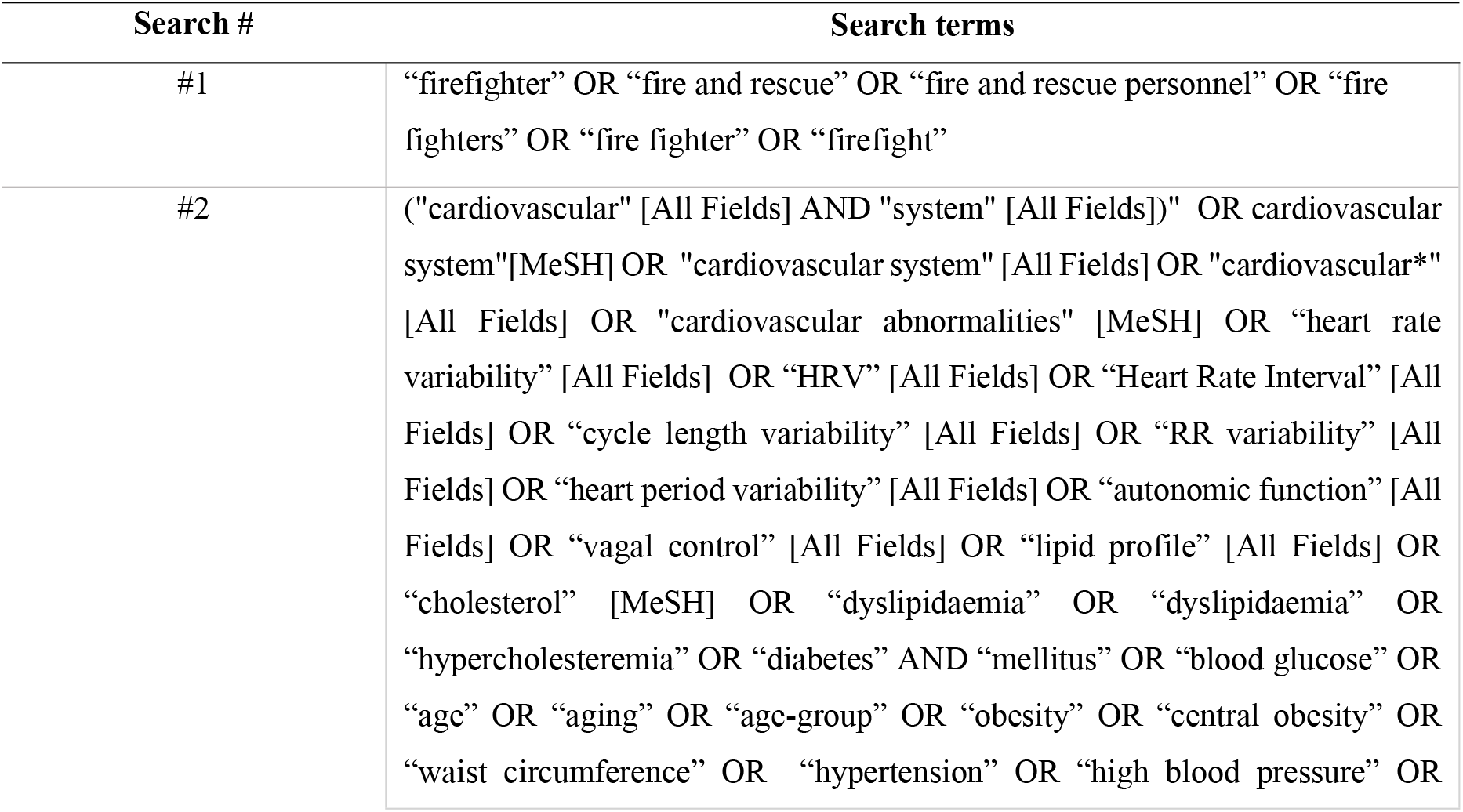
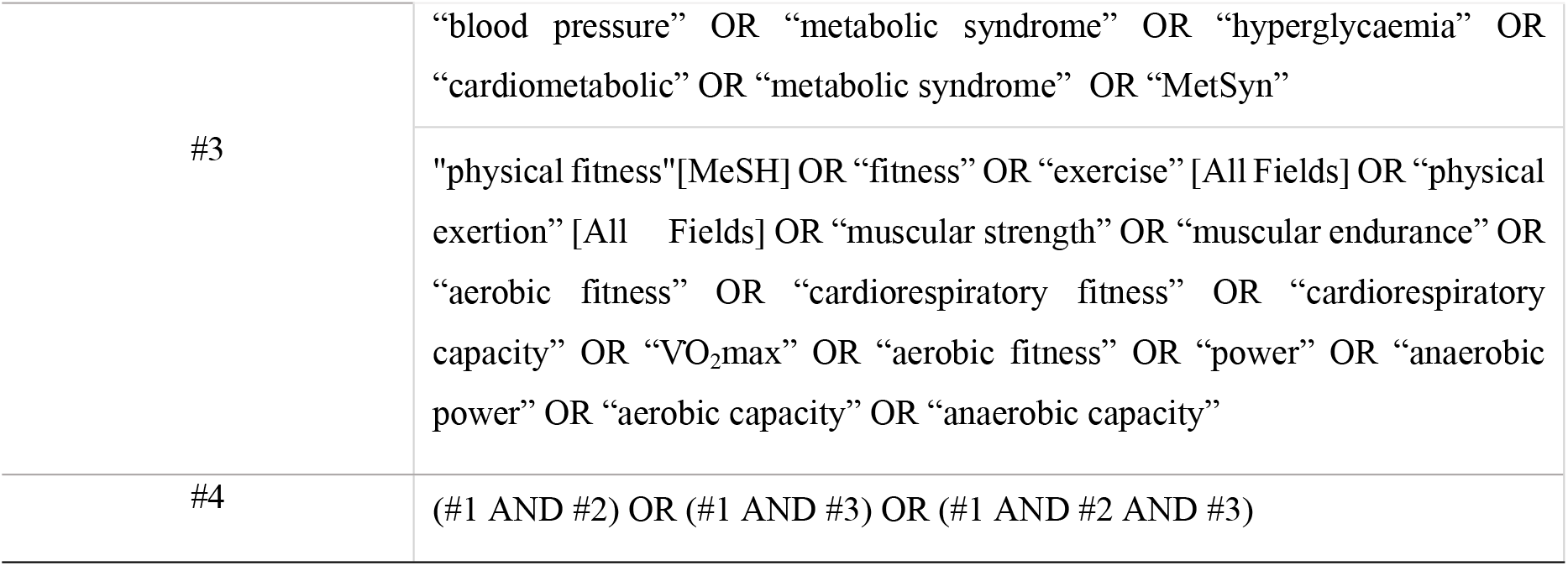
Search strategy developed in PubMed.

### Study Selection

Initially, the electronic database search will be performed to acquire all eligible research papers. All studies that meet the inclusion criteria will be selected for full-text screening. Authors will be contacted in the instance where data is missing or where a full-text is not available. Subsequently, the full-text articles will be assessed independently by the two principal reviewers (JR and RN), using the Rayyan® intelligent systematic review (RIS) tool [33]. When screening and categorizing the studies, three categories will be used, specifically, included, excluded and unsure. Where uncertainties in study inclusion or study grading, the dispute will be discussed between the two reviewers, and in the event of disagreement, the adjudicator (LL) will resolve the dispute.

#### Selection and Screening of Articles

Firstly, all relevant, pre-selected databases will be searched to identify and, initially, screen the titles and abstracts of potential studies for eligibility. Secondly, the search outputs will be downloaded and compiled in a reference software, namely, Zotero® version 6.0.9 and thirdly, the reference software will be used to remove duplicate studies. Fourthly, the studies remaining after the removal of duplicates, will be screened against the inclusion criteria to determine the final studies for inclusion in the review and meta-analysis. Next, data will be extracted from the included studies using a standard data extraction form. Lastly, studies included into the meta-analysis will be analysed and interpreted using Review Manager 5.3[34] and MedCalc® statistical software Ltd.

### Data Management

All data that will be extracted from the initially selected studies will be captured using an initial electronic researcher-generated data extraction form (Appendix 2), by the two principal reviewers, in order to retrieve the key characteristics of each study. Thereafter, a detailed second data extraction form (Appendix 3) will be used to capture information of the included studies. The study data that will be extracted will include the general details of the study, such as the study title, authors, publication date, study design and the country of origin, the exposures evaluated, and the outcome measured. Thereafter, characteristics of the included studies will be collected, such as the sample size and method of sampling, participants details (age, height, weight, body mass index, cardiorespiratory fitness, gender). Lastly, the details of exposures and the outcome variables will be extracted, i.e., the research paper must report on at least one of the CVD risk factors under investigation in relation to cardiorespiratory fitness.

#### Critical Appraisal of Included Studies

Two critical appraisal tools will be used for this systematic review and meta-analysis, namely, The Critical Appraisal Skills Programme (CASP) toolkit (Middle Way, Oxford, UK) and the Appraisal Tool for Cross Sectional Studies (AXIS) checklist [35] (Table 2) (https://casp-uk.net/casp-tools-checklists/) to conduct the methodological quality assessment of each study that was included. The CASP toolkit and the AXIS toolkit was shown to be a reliable and valid tool for assessing the quality of cross-sectional studies [35,36]. For the grading of each study, where questions can be answered dichotomously, an article will be awarded a yes, denoted as a “□”, and when scored a no, an “×” will be denoted. For questions that require written grading, all attempts will be made to adapt the question to a dichotomous scale, and if a rating is not possible, the question will be excluded from the checklist. For questions that are not relevant, an “na” will be given.

### Data Synthesis and Meta-Analysis

Data synthesis will commence once all relevant details of each study has been captured. Initially, a systematic synthesis of the results will be used to integrate the findings, narratively, from the various studies. This method will be used initially as it allows the researcher to identify, evaluate, and summarize study findings that are similar across all relevant studies [37–39]. Thereafter, data will be captured, quantitatively, and analysed using a meta-analysis.

#### Meta-Analysis

Where there is sufficient reporting and the data permits, a meta-analysis will be conducted. For each study, quantitative data will be pooled to determine the pooled effect estimate [40]. For dichotomous data, the odds ratio (OR) and risk ratio (RR) will be generated, whereas for continuous data, the mean difference (MD) and standardized mean difference (SMD) of estimation will be used to estimate the effect of each cardiovascular disease risk factor on cardiorespiratory fitness of firefighters [41]. The data will be imported and analysed using the Review Manager 5.3 [34,39,41,42], using the inverse method of meta-analysis [40]. In addition, a meta-analysis of correlations will be performed to determine the relationship between continuous CVD risk factors and cardiorespiratory fitness of firefighters. To perform the analysis, the MedCalc® statistical software Ltd. (version 20.104) will be preferred [40]. The meta-analysis will involve using the original sample sizes and r values to generate the pooled r values between each cardiovascular risk factor in relation to cardiorespiratory fitness [40]. Using the Fisher’s r to z transformation the original r values will be converted to a common test metric [40]:

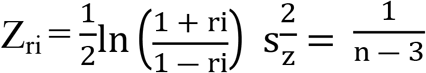

The following will be used to indicate the strength of correlation [40]:

1. Very high correlation: 0.90 to 1.00 (−0.90 to −1.00).
2. High correlation: 0.70 to 0.90 (−0.70 to −0.90).
3. Moderate correlation: 0.50 to 0.70 (−0.50 to −0.70).
4. Low correlation: 0.30 to 0.50 (−0.30 to −0.50).
5. Negligible correlation: 0.00 to 0.30 (−0.00 to −0.30).

#### Assessment of Heterogeneity

Heterogeneity will be evaluated, initially, through visual inspection of the forest plot, to judge the extent of overlap. In addition, to quantitively assess the heterogeneity, the Chi-square test and the I^2^ test will be used [60]. The following will be used to explain I^2^ statistics: 1) 0% to 30% not be important; 2) 31% to 60% moderate heterogeneity; 3) 61% to 80% substantial heterogeneity; and 4) 81% to 100% considerable heterogeneity [40,42].

In the instance where substantial and considerable heterogeneity is found, potential reasons will be determined by performing a subgroup analysis. A meta-analysis will be favoured by a low degree of heterogeneity [60]. Where significant heterogeneity is present, then a descriptive analysis of the results will be presented. If homogeneity is found between the studies, then a fixed-effects model will be preferred, and if heterogeneity is present, a random-effects model will be preferred [60].

#### Subgroup Analysis and Investigation of Heterogeneity

The following characteristics are expected to introduce significant clinical heterogeneity, i.e., firefighters’ age, gender, experience, methods of measuring CVD risk factors, type of testing procedure used to assess cardiorespiratory fitness, and plan to implement subgroup analysis on these variables, if possible. Although all exposures and outcomes will be measured using standardized instruments and protocols, the protocols preferred by each study may be different. This will necessitate the comparison and conversion of certain variables to produce analogous findings for comparison. The authors will use Review Manager for the subgroup analysis [39,41,42], and MedCalc® statistical software Ltd.[40] if adequate numbers of included studies are available.

#### Publication Bias

Publication bias will be assessed visually using the Begg’s funnel chart. Thereafter, quantitative analysis will be conducted to assess the possibility of publication bias using the Egger’s test and Begg’s test [40].

#### Presenting and Reporting of Results

The results generated will be presented using a combination of figures, graphs, and tables. The presentation will include the method involved in study selection, exclusion and inclusion, using the PRISMA guidelines [32]. In addition, forest plots, funnel charts and summary tables will be created to present the data.

## DISCUSSION

To the best of the authors’ knowledge, no conclusive evidence currently exists on the effect of CVD risk factors and cardiorespiratory fitness of firefighters. This systematic review arose from the global cardiovascular health concerns that firefighters face, and how the development and progression of CVD risk factors effect cardiorespiratory fitness in firefighters. Cardiorespiratory has been shown to be an important factor in optimal occupational performance in firefighters, but understudied [21,43–45]. The results of this review is expected to make a significant contribution to the international scientific literature on the effect that CVD risk factors will have on cardiorespiratory fitness. In addition, the results of this review will assist policy makers in implementing policy changes, particularly in developing countries, such as South Africa, but also globally, to promote the wellbeing, health, and career longevity of firefighters. Moreover, the planned review will support researchers design original studies relating to this issue and, assist in identifying gaps in the research for further studies.

As with all research, there are inherent limitations in the current systematic review and meta-analysis protocol when interpreting the findings. One limitation of this study is that only English studies will be used, which may result in the exclusion of potentially relevant studies. Another limitation is that there may be considerable heterogeneity between studies, due to differences in study population age, gender and the method used to assess cardiorespiratory fitness. There are, however, several strengths of this study, one of which being that this review will be guided by reference methodologies to ensure that a high standard of quality is maintained throughout the review process. The exclusion of a publication date limit is a strength of this review, removing the possibility of high-quality studies being excluded due to falling outside the publication limit. Lastly, the inclusion of all study designs, using various study methodologies, in this review.

### Ethics and Dissemination

This study has been registered onto PROSPERO (CRD42022330510) and has been granted ethical clearance by the University of the Western Cape (BM21/10/9). Because a systematic review and meta-analysis will be conducted, there will be no direct engagement with human participants. Freely accessible and published data will be used in the study and, therefore, no confidentiality or ethical procedures need to be considered for this review [46]. The information gathered will be presented to local firefighter organisations and health departments, at conferences and in webinars. Furthermore, the systematic review and meta-analysis will be published in an accredited peer-reviewed journal. Lastly, this protocol will form a chapter of a doctoral thesis.

## Data Availability

No datasets were generated or analysed during the current study. All relevant data from this study will be made available upon study completion.

## AUTHOR CONTRIBUTIONS

**Conceptualisation**: Jaron Ras

**Data curation:** Jaron Ras

**Formal analysis:** Jaron Ras

**Funding acquisition:** Jaron Ras

**Investigation:** Jaron Ras

**Methodology:** Jaron Ras, Elpidofos Soteriades, Denise Smith, Andre Kengne and Lloyd Leach

**Project administration:** Jaron Ras, Elpidofos Soteriades, Denise Smith, Andre Kengne and Lloyd Leach

**Resources:** Jaron Ras, Elpidofos Soteriades, Denise Smith, Andre Kengne and Lloyd Leach

**Supervision:** Elpidofos Soteriades, Denise Smith, Andre Kengne and Lloyd Leach

**Writing – original draft preparation:** Jaron Ras

**Writing – review and editing:** Elpidofos Soteriades, Denise Smith, Andre Kengne and Lloyd Leach

**Protocol Registration**

Details of the protocol for this systematic review were registered on PROSPERO (CRD42022330510) and can be accessed at https://www.crd.york.ac.uk/prospero/display_record.php?RecordID=330510

